# Targeting students of nonhealth academic fields for basic life support: they need to know why, what and how to do CPR

**DOI:** 10.1101/2024.03.04.24303753

**Authors:** PN Sugimoto, GB Gouvêa, IC Salles, HB Carvalho, P Aikawa, LMTA Azi, LFF Silva, M Macchione, F Semeraro, A Lockey, RT Greif, MJC Carmona, BW Böttiger, NK Nakagawa

**Affiliations:** Education, Assessment and Intervention in Cardiovascular Group, Faculdade de Medicina da Universidade de São Paulo, São Paulo, Brazil; Preventive Department, Faculdade de Medicina da Universidade de São Paulo, São Paulo, Brazil; Institute of Biological Sciences, Federal University of Rio Grande, Rio Grande do Sul, Brazil; Federal University of Bahia, Bahia, Brazil; Department of Anesthesia, Intensive Care and Prehospital Emergency, Maggiore Hospital Carlo Alberto Pizzardi, Italy; Calderdale and Huddersfield NHS Trust, Halifax, United Kingdom; School of Medicine, Sigmund Freud University Vienna, Austria; and University of Bern, Switzerland; Anesthesiology Discipline, Faculdade de Medicina da Universidade de São Paulo, Brazil; University of Cologne, Department of Anaesthesiology and Intensive Care Medicine, University Hospital, Medical Faculty, Germany

**Keywords:** Sudden cardiac arrest, Cardiopulmonary resuscitation, Basic life support, Acute myocardial infarction, Bystander, Automated External Defibrillator

## Abstract

Education in basic life support is widely proposed to increase survival and quality-of-life in out-of-hospital sudden cardiac arrest. We aimed to assess knowledge, skills and attitudes regarding acute myocardial infarction and sudden cardiac arrest among university students of all fields of knowledge.

**Methods:** The local Ethical Research Committee approved this cross-sectional study. An electronic survey “KIDS SAVE LIVES BRAZIL” was sent to 58,862 students of 82 disciplines in three universities, aged ≥ 18 years. The survey covered three categories: knowledge, skills, and attitude. Each category was graded between 0 and 10 points (the highest).

**Results:** Among university students, 4,803 undergraduates (8.2 %) answered the survey, and were divided in three groups of disciplines: medicine (219, ∼21.7 years, 38% male), other-health-care (n=1,058; ∼22.9 years; 36% male), and nonhealth-care (n=3,526; ∼22.9 years; 35% male). All three groups showed significant differences between them (p<0.001). The nonhealth-care compared with medicine and other-health-care group showed, respectively, the lowest (p<0.001) median scores (25-75%) in knowledge [4.0 (0.0-9.3), 4.0 (4.0-8.0), and 4.0 (4.0-4.7)], skills [2.4 (1.2-3.3), 6.4 (4.0-8.3), 4.0 (2.4-6.2], and attitude [5.9 (5.9-6.8), 7.3 (5.9-7.3), and 7.3 (5.9-7.3)].

**Conclusion:** University students have the willingness to help victims suffering from acute myocardial infarction or sustaining sudden cardiac arrest. However, nonhealth-care students markedly lack knowledge and skills to perform cardiopulmonary resuscitation and automated external defibrillation. Our findings reveal a stark difference in basic life support competencies between students in health-care related fields and those in nonhealthy-care fields, emphasizing the need for universal basic life support training.

**KEY MESSAGES:** 1. Our findings reveal a stark difference in basic life support competencies between students in health-care related fields and those in nonhealth fields, emphasizing the need for universal basic life support training.

2. An action for curriculum modification to include basic life support training for all students is timely and practical, given the global burden of heart disease and the proven benefits of early intervention in sudden cardiac arrest cases.

3. Our study contributes significantly to the ongoing discussion about public health education and the role of nonhealth professionals in emergency medical response. It may serve as a catalyst for policy changes within educational institutions and among healthcare policymakers, aiming to create a more resilient and responsive community in the face of out-of-hospital medical emergencies.

## INTRODUCTION

Acute myocardial infarction (AMI) can be a cause of arrhythmia and out-of-hospital sudden cardiac arrest (OHCA) in adults.^1^ Both, AMI and OHCA, are major public health burdens with high incidence, high morbidity and high mortality rates worldwide.^2, 3^ Survival rates after OHCA are globally low (<10%), and are proportional to the rate of people trained in early high-quality cardiopulmonary resuscitation (CPR), and adequate use of an automated external defibrillator (AED).^3-5^

In the last two decades, studies showed that geographical region, racial distributions, bystanders aged > 30 years, and low income are associated with lower probabilities of bystanders initiating CPR in OHCA.^3, 6, 7^ Recently, education for laypeople in CPR and AED use have been widely proposed since childhood to improve quality of life^8^ and to increase survival after OHCA by the World Health Organization,^9^ and the International Liaison Committee on Resuscitation.^10-11^

BLS training follows the current international guidelines, and young adults have favorable cognitive and physical characteristics (larger than 1.5 meter, more than 50 kg or body mass index higher than 22 kg.m.-2 to adequately perform CPR and to use AED.^12, 13^ Undergraduate students of medicine,^14-16^ nursing, physiotherapy, and other academic health care disciplines^17^ have BLS included in their curriculum. However, little is known on nonhealth-care university students competencies on BLS.

This study is based on an electronic survey and aimed to investigate self-rated knowledge, skills and attitude on AMI and OHCA among university students of all disciplines divided in medicine, other health-care disciplines and nonhealth-care disciplines.

## METHODS

This is a cross-sectional observational study of the KIDS SAVE LIVES BRAZIL project with approval of the Ethical Committee of the Faculty of Medicine, São Paulo University, Brazil (CAAE: 25218819.0.0000.0065). We recruited students from 82 different higher education disciplines, aged ≥ 18 years, both sexes, from three public universities of Brazil (University of São Paulo, State University of Campinas, and Federal University of Rio Grande). An electronic survey was sent only once to all students by the Rectory of the University, from March 2020 to February 2022. We included students who agreed with the research written informed consent, and who answered the survey until April 2022. We excluded those students who did not provide information on age or university discipline. Students were allocated in the following three groups of disciplines: medicine, other-health-care and nonhealth-care.

We performed a sample size calculation by estimating a population proportion of 15% that would grade at least five in knowledge with a specified absolute proportion of 1%, that resulted in 4,898 students. The formula was^18^

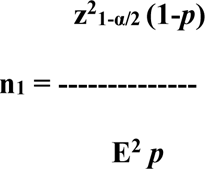

Where: n_1_ is the estimated sample size; E is the assumed level of confidence of 95%; z is the z curve area for different levels of significance; and p is the expected proportion for the event.

### Electronic Questionnaire

Several aspects of the present survey were based on our previous work^19^ which aimed to understand the scenario in primary, secondary and high schools on key principles of KIDS SAVE LIVES BRAZIL. We created an electronic survey in the Google form platform, that was available at the University. Each student performed a self-rating answer. It is a 25-item multiple choice online questionnaire covering three categories: knowledge (5 items), skills (8 items), and attitude (12 items) towards OHCA and AMI. Each category (knowledge, skills and attitude) was graded between zero and 10 (the highest grade). The questions included in each category are in the Supplementary Material.

### Statistical Analysis

We used the Statistical Software STATA for Windows (Stata Corp LLC. Texas, USA), version 24.

We applied the Shapiro-Wilk Test for normal distribution analysis of age and grades in each category (knowledge, skills and attitude).

Data of the three groups of disciplines: medicine, other-health-care and nonhealth-care are presented as means (± standard deviation) if continuous data show normal distributions, and they were analyzed using the ANOVA Test. Data with non-normal distributions are presented as medians (25-75% quartiles), and were analyzed using the Kruskal-Wallis Test. When appropriate, a multiple-comparison paired analysis was performed using the Dunńs Test.

Categorical variables are presented as numbers and proportions. The categorical variables were sex and grades above (yes/no) the median score of all students of all disciplines in each category (knowledge, skills and attitude). We used Chi-Squared Test to analyze data among the three groups.

A p-value < 0.05 was considered statistically significant.

## RESULTS

We sent the electronic survey to 58,862 undergraduates of 82 disciplines from three public Universities (Fig. 1). A total of 4,803 students (8.2%) answered the survey: 5% of the students was studying medicine (n=219), 22% was studying other-health-care disciplines (n=1,058), and 73% was studying nonhealth-care disciplines (n=3,526).

**Figure 1.**
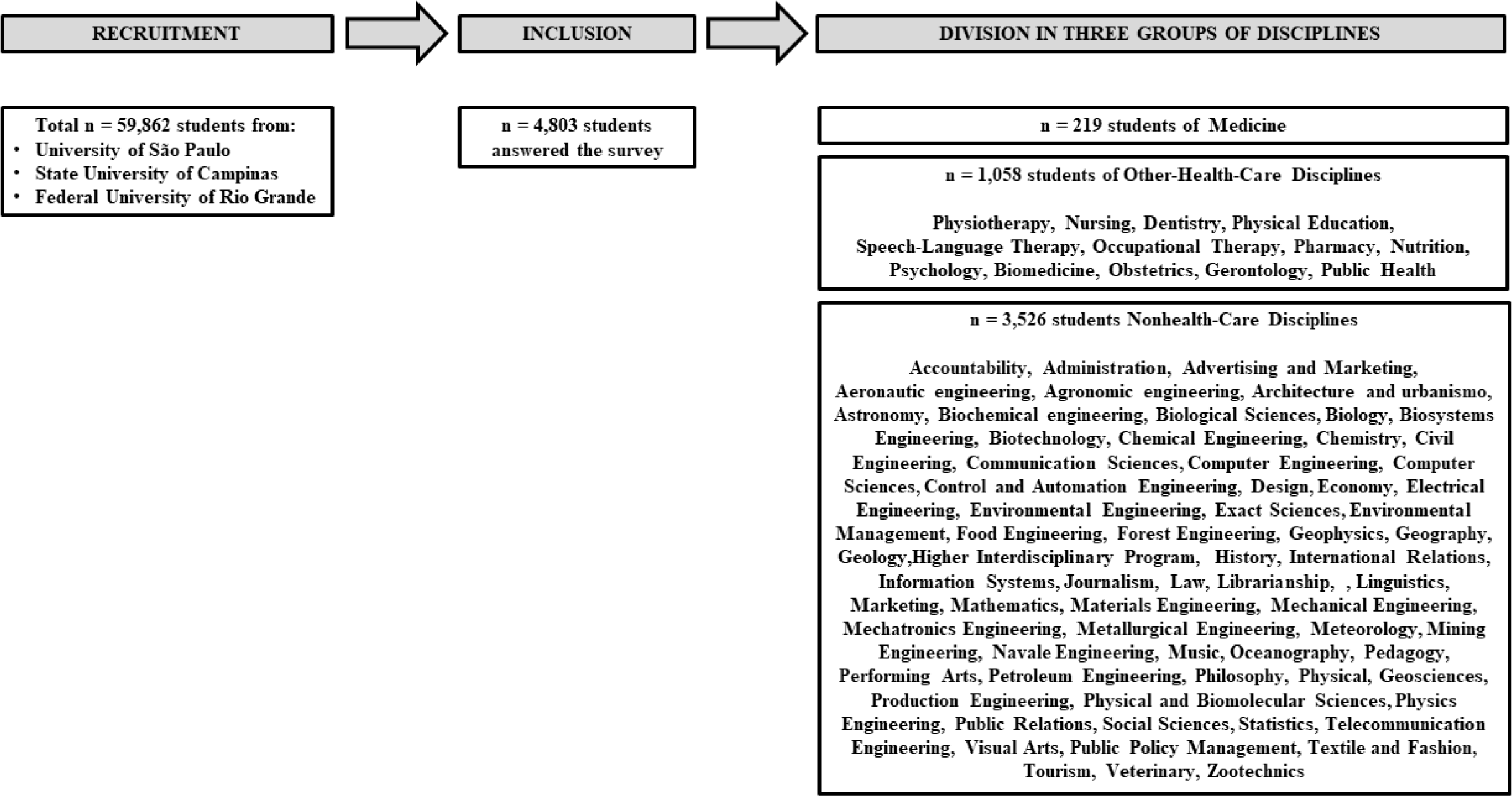
The cross-over study design using an electronic survey sent to all students of all years of three universities. Responders of 82 disciplines were divided in three groups of disciplines: medicine, other-health-care, and nonhealth-care

Students in medicine were younger than students of the other-health-care and nonhealth-care disciplines (Table 1). Female students were prevalent in this study. However, sex proportions were similar between the three groups of disciplines.

**Table 1.**
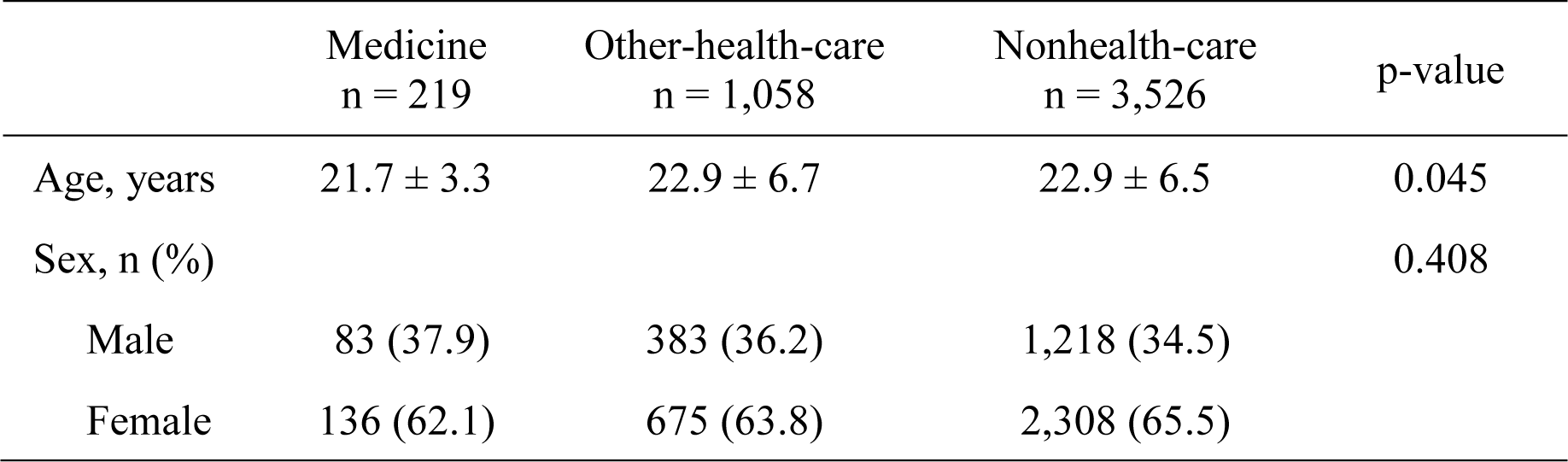
Demographic characteristics are presented as mean values and standard deviations or numbers of subjects and proportions. Comparisons between groups of disciplines (medicine, other-health-care, and nonhealth-care) were performed using the ANOVA test or the Chi-Squared Test, when appropriate.

Grades in the categories knowledge, skills and attitude are presented in Figure 2. The median score (25-75% quartiles) of all students of all disciplines in knowledge was 3.2 points (0-4.0). After post-hoc analysis with the Dunńs Test, the nonhealth-care students showed the lowest (p<0.001) median score (25-75% quartiles) of 4.0 (0-9.3) than medical students with a median score of 4.0 (4.0-8.0) and, the other-health-care students with a median score of 4.0 points (4.0-4.7).

**Figure 2.**
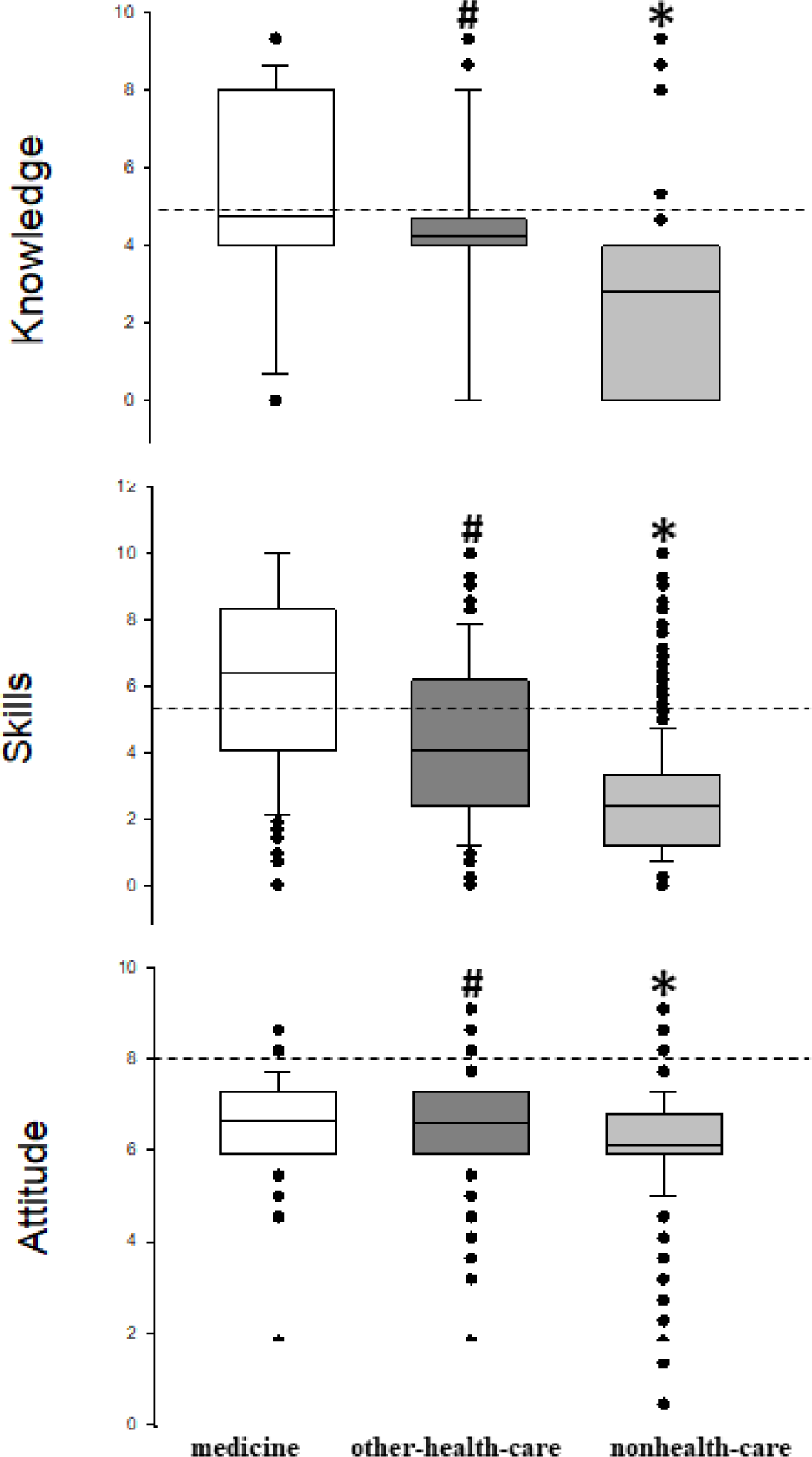
Box-plot of self-reported grades in three categories (knowledge, skills and attitude) by university students of medicine (┌), other-health-care disciplines (▄) and nonhealth-care disciplines (▄). Dashed lines show the median values of all students in each category. Significant differences were found between groups using the Kruskall-Wallis Test with post-hoc analysis (Dunńs Test) (* p <0.001 *vs* other groups; # p<0.001 *vs* medicine)

The median score (25-75% quartiles) of perceived skills reported by all students of all disciplines was 3.1 (1.7-4.1). The three disciplines showed significant different median scores between them in skills (p<0.001). Medical students had the highest score of 6.4 (4.0-8.3). The other-health-care students had 4.0 (2.4-6.2). The nonhealth-care group obtained the lowest score of 2.4 (1.2-3.3)(Figure 2).

The median score (25-75% quartiles) in attitude reported by all students of all disciplines was 6.2 (5.9-7.3) and there were significant differences between all three groups. The nonhealth-care university students had lower (p<0.001) median score of 6.1 points (5.9-6.8) than the other two groups (Figure 2). The medicine group obtained similar median scores (25-75%) of 6.6 (5.9-7.3) as the other-health-care disciplines, however, median was greater. Among 4,803 respondents, 98% (n=4,708) showed the willingness to help people in SCA. Similarly, 98% respondents (n=4,747) wished to have practical BLS course at school.

**Table 2.** Numbers and proportions of students with grades above the median score of all students of all disciplines in each category (knowledge, skills and attitude). Comparisons between groups of disciplines (medicine, other-health-care, and nonhealth-care) were performed using the Chi-Squared Test (*, *vs* the other groups; #, *vs* medicine)

|  | Medicine<br>n = 219 | Other-health-care<br>n = 1,058 | Nonhealth-care n<br>= 3,526 | p-value |
| --- | --- | --- | --- | --- |
| Knowledge | 65 (29.7) | 295 (27.9) | 160 (4.5)* | <0.001 |
| Skills | 185 (84.5) | 687 (64.9)# | 1,298 (36.8)* | <0.001 |
| Attitude | 150 (68.5) | 594 (56.1) | 1,088 (30.9)* | <0.001 |

In the analysis of the proportions of students that graded above the median score in each category (Table 2), we found that nonhealth-care disciplines had reduced proportions of students compared with the medical and other-health-care disciplines in two categories knowledge and skills.

## DISCUSSION

To our knowledge, this is the first study investigating self-rated knowledge, skills and attitude in SCA and AMI among students of 82 different disciplines in multiple higher education institutions of a lower resource country. Students (aged about 23 years and 65% female) were grouped in three groups of disciplines: Medicine, Other-health-care and Nonhealth-care. The present study reveals that independently of health-care or nonhealth-care related fields, we found significant differences in BLS knowledge, skills and attitude between students of health-care related disciplines and nonhealth-care disciplines. Additionally, the great majority (98%) of the university students of all disciplines reported the willingness to help victims of SCA. Taken together, our study may emphasize the need for universal BLS training.

We found significant differences between groups in knowledge, skills and attitude. As expected, in the present study, medical students rated the highest scores in knowledge, skills and attitude facing SCA and AMI. The year of medical school and type of BLS training vary among countries and geographical regions,^20^ but most studies in the literature have been performed with medical students in their final year of school.^21^ Students in the 6^th^ year of medical school may achieve higher grades as they are supposed to have had several opportunities along the six years curriculum for competencies (knowledge, skills and attitude) development to manage and deal with SCA and AMI. However, by including students of all six years medical school, we intended to overcome this volunteer bias.

Students of other-health-care academic disciplines (all years of university) were also investigated. They rated lower median grades in knowledge, skills and attitude in SCA and AMI than medical students. This result can be explained by the fact that only few disciplines (dentistry, nursing, and physiotherapy) also have BLS training, and other contents on AMI, and SCA in their curriculum.^17^ Other-health-care disciplines included in this study, such as physical education, speech therapy and occupational therapy did not have BLS practical training in their curriculum.

Interesting findings are also in attitude category. For instance, median scores with IQR were very similar between medicine and other-health-care disciplines, however, results were significantly different because the Dunńs Test takes into account the number of students in each group.^22^ Other interesting aspects found in the present study were that, independently of health-care related or nonhealth-care disciplines, the great majority of students (98%) showed the willingness to help people in medical emergencies as wished to have BLS course at school. However, students of nonhealth-care disciplines have nothing related to BLS theory or practice on why, what, and how to respond to save a life in their academic curricula at the university.^20^ University students are, in general, young adults with favorable cognitive and physical characteristics to perform BLS.^8^ Then, it is of note that the nonhealth-care students compose an important larger target group of the society to develop competencies as first responders in out-of-hospital emergencies. Taken together, these results raise the need of curriculum modification by adding BLS training to students of all academic disciplines at higher education. University students should know why, what and how to respond in out-of-hospital emergencies, taking first actions in SCA and AMI until the emergency medical service arrival. Each individual may have his/her own way of learning, and a great variety of educational modalities can be used for this purpose.^7, 8^

Our study has limitations. The electronic survey on emergency situations was sent to approximately 59,000 students of three public universities. With only 8.2% of respondents, one may argue that our results may not be extrapolated to the broader population. Indeed, the number of respondents (4,803) was lower than that estimated (n=4,898). However, we studied several segments among students by inviting all students of the three public universities to participate in the survey. There is a national policy that reserves 50% of the total entry places in public universities for students that passed in the exams with low-income, and/or blacks, and/or indians. In this context, we speculate that our sample of students in higher education would have a relative wide demographic and different backgrounds (low, middle and high social-economic status, different races, different school years, etc) across multiple institutions. Another aspect is that this survey was not validated. We dońt know if it measures what is supposed to measure, and more studies are needed to assess its validity to generate similar results if repeated.

Another limitation was that we used a self-rated and directive electronic survey, focusing on BLS skills and attitude, with no practical observations or objective measures. This is a study based on a self-rating online questionnaire that may induce bias as respondents tend to over-report normative behavior, and directive survey may increase the likelihood of respondents to report certain behavior that would appear socially agreeable.^23^ However, in this context, some aspects may help to overcome this bias, such as, the respondent identity was covered, the survey was not sent by a specific discipline, or related to a specific academic aspect.

Taken together, our findings could contribute to the ongoing discussion about public health education and the role of nonhealth-care students and professionals in emergency medical response. Our study calls for curriculum modification to include BLS training for all students of all disciplines in higher education. Nowadays, this can be considered timely and practical, given the global burden of heart disease and the proven benefits of early intervention in SCA cases. It may serve as part of a worldwide catalyst for policy changes within educational institutions and among healthcare policymakers, aiming to create a more resilient and responsive community in the face of out-of-hospital medical emergencies.

In conclusion, university students have the willingness to help victims suffering from acute myocardial infarction or sustaining sudden cardiac arrest. However, nonhealth-care students markedly lack knowledge and skills to perform cardiopulmonary resuscitation and automated external defibrillation. Our findings reveal a stark difference in basic life support competencies between students in health-care related fields and those in nonhealthy-care fields, emphasizing the need for universal basic life support training.

## Data Availability

Data will be available after 6 to 12 months of the publication with direct request to the authors.

## ACKNOWLEDGEMENTS

The authors would like to thank the children and adolescents of the public schools that participated in this study, all members of the KIDS SAVE LIVES BRAZIL Project, the European Resuscitation Council, and Prof. Dr. Ana Paula Boaventura for helping us to recruit students from UNICAMP.

## AUTHOR CONTRIBUTIONS

CONCEPTUALIZATION: P.N.S., G.B.G., I.C.S., H.B.C., P.A., M.J.C.C., N.K.N.

METHODOLOGY: H.B.C., P.A., L.F.F.S., M.M., M.J.C.C., N.K.N.

SOFTWARE: H.B.C.

VALIDATION: N.K.N., H.B.C.

FORMAL ANALYSIS: P.N.S., G.B.G., I.C.S., N.K.N., H.B.C.

INVESTIGATION: P.N.S., G.B.G., I.C.S., N.K.N.

RESOURCES: H.B.C., N.K.N.

DATA CURATION: P.N.S., G.B.G., I.C.S., H.B.C., N.K.N.

WRITING – ORIGINAL DRAFT PREPARATION: P.N.S., G.B.G., I.C.S., H.B.C., P.A., M.M., M.J.C.C., N.K.N.

WRITING – REVIEW & EDITING: H.B.C., P.A., A.P.B., L.M.T.A.A., L.F.F.S., M.M., F.S., A.L., K.G.M., R.G., M.J.C.C., B.W.B., N.K.N.

VISUALIZATION: P.N.S., G.B.G., I.C.S., H.B.C., P.A., M.M., M.J.C.C., N.K.N.

SUPERVISION: H.B.C., P.A., M.J.C.C., N.K.N.

PROJECT ADMINISTRATION: M.J.C.C., N.K.N.

FUNDING ACQUISITION: N.K.N.

